# Risk Factors for Non-Communicable Diseases among Bangladeshi Adults: An Application of Generalized Linear Mixed Model on Multilevel Demographic and Health Survey Data

**DOI:** 10.1101/2023.12.07.23299668

**Authors:** Kazi Sabbir Ahmad Nahin, Tabita Jannatul

## Abstract

**Objective:** The research effort addresses the impact of non-communicable diseases (NCDs), particularly diabetes mellitus (DM) and hypertension (HTN), on Bangladesh, a lower-middle-income country. Due to their higher incidences and associated risks, DM and HTN present substantial concerns. The paper clarifies the need for specific public health initiatives and emphasizes the socioeconomic and lifestyle-related factors of NCDs.

**Methods:** A mixed-model technique is employed to conduct a multivariate analysis of the cross sectional data from the Bangladesh Demographic and Health Survey (BDHS) 2017-18, with the aim of identifying possible risk factors. We have two outcome variables under consideration in the study, namely Diabetes, and Hypertension, each having binary categories. The adjusted odds ratios (AORs) in addition to their corresponding p-values and 95% confidence intervals (CIs) to assess and evaluate the relative strength of covariates.

**Results:** Those with hypertension have a 28% (OR = 1.28, 95% CI: 1.14, 1.43) higher risk of developing diabetes. Likewise, individuals with diabetes have a 24% (OR = 1.24, 95% CI: 1.11, 1.39) increased probability of developing hypertension. People over 40 years old are 66.4% more likely to have diabetes. Also, rich people are more likely to be diabetic and hypertensive.

**Conclusion:** The prevalence of diabetes is significantly elevated among individuals aged 40 years and older, particularly among those who have hypertension, are overweight, and possess a higher socioeconomic status. In contrast, there is a notable increase in the likelihood of developing hypertension among male individuals who are diabetic, above the age of 40, who already have hypertension, are overweight and have a higher socioeconomic class. There is a good chance that presenting these risk factors to policymakers could contribute to the amelioration of the NCDs crisis in Bangladesh.

## Background

[1] Non-communicable diseases (NCDs) kill 41 million people each year, responsible for more deaths than all other causes combined, contributing to 71% of all deaths worldwide. [2] NCDs comprised not only 7 of the top 10 main causes of mortality, but they also accounted for 74% of all deaths worldwide in 2019. [1] Each year, around 15 million individuals between the ages of 30 and 69 years die of an NCD; 85% of these “premature” deaths (30 to 70 years) occur in lower-and-middle income countries(LMICs). [3,4] Bangladesh as an LMIC, is expected to suffer from NCDs at a proportion of 59% of total deaths-886,000 deaths per year.

[5] NCDs impact millions of people throughout the globe, and they affect individuals at all phases of life, from infancy to old age. Beyond the negative impact on people’s lives, NCDs pose a serious threat to public health, as well as to the nation’s economic and social progress.

[6,7,8,9,10] Following the recent series of publications of the Lancet Taskforce on NCDs and Economics, it has been observed that there is a substantial link between economic development and the supervision of NCDs. Poverty adds to the detrimental consequences of NCDs. [11] NCDs impose a considerable financial burden on healthcare systems and the welfare of countries, which is likely to grow in the future.

[12,13,14] As it is anticipated that the number of NCD fatalities would increase from 41 million to 52 million by 2030, and given the adverse repercussions that NCDs have on people as well as the global community, the prevalence of NCDs is unquestionably a barrier to accomplishing the Sustainable Development Goal (SDG) target 3.4, which aims to “By 2030, reduce by one-third premature mortality from non-communicable diseases through prevention and treatment and promote mental health and well-being”. Even while the good news is that NCDs are receiving greater attention in developed countries, we must also address the underlying factors that are contributing to the growth of NCDs in developing countries.

[15] World Health Organization(WHO) identified four global risk factors for the rise of NCDs: tobacco use, physical inactivity, the harmful use of alcohol, and unhealthy diets. [1,16] Subsequently, these cause four metabolic changes: elevated blood pressure (hypertension), obesity, elevated blood glucose(hyperglycemia), and elevated blood lipids(hyperlipidemia). [16,17] Since about 90% of Bangladesh’s population is Muslim, alcohol usage is deemed to be a minor issue due to religious restrictions. [16] However, recent years have shown a rise in other factors. [18,19,20,21,22] People in Bangladesh are demanding prompt response since the burden of diabetes mellitus (DM), hypertension (HTN), and other major contributors of NCDs is mounting amidst initiatives to improve healthcare and well-being.

[23] Diabetes takes the life of one person every five seconds. [24] As of 2017, there are an estimated 451 million individuals between the ages of 18 and 99 who live with diabetes. By 2045, these numbers are predicted to rise to 693 million people. Besides 49.7% of diabetic patients are undiagnosed, according to the study. [25] The prevalence of diabetes has been growing substantially over the last three decades, with the fastest increases occurring in LMICs. [26] LMICs account for around 80% of all diabetic patients, with the vast majority of these individuals remaining undiagnosed for years. [27] An estimated 1.5 million people died in 2019 as a direct result of diabetes, making it the ninth-highest cause of death worldwide in 2019. [28] By 2030, the incidence of diabetes in the South Asian region is expected to rise by more than 150%, making it a major public health issue. [29] Following a scoping review, it was discovered that both urban and rural communities in Bangladesh were experiencing an increase in the prevalence of type 2 diabetes mellitus (T2DM). [30] According to the findings of a systematic review and meta-analysis, the proportion of DM in Bangladesh was 7.8 (95% CI:6.4–9.3). [31] As per the WHO, DM affects 12.88 million (8% of total population) individuals in Bangladesh, and it is responsible for 3% of all fatalities.

[32] Worldwide, HTN has become one of the leading causes of “premature” deaths, a “silent killer” according to the WHO, considering fewer than one in five individuals have it under control. [33] It is estimated that HTN is responsible for the deaths of 9.4 million people annually throughout the globe, which is roughly the same number as the total deaths due to infectious diseases. [34] The prevalence of individuals with hypertension is anticipated to increase by approximately 60% to 1.56 billion (95% CI:1.54–1.58 billion) by 2025. [32,35] Recent findings reveal that LMICs are responsible for two-thirds of the world’s HTN burden, contrasting with the general view that the condition is more prevalent in wealthy nations. [36] South Asian Association for Regional Cooperation (SAARC) nations have quite a high prevalence of HTN that is above the global norm, as shown in a meta-analysis and systematic evaluation of data. [37] HTN prevalence seems to be on the upswing in Bangladesh, according to the latest research, with the percentage reaching 20%.

[38,39,40,41] The key contributions towards the development of NCDs contain numerous socioeconomic, sociodemographic, and lifestyle-related risk factors. [42,43,44] Both sexes in East and South Asia are seeing an upsurge in BMI (general obesity index), which has been linked to an increased risk of developing DM. [45] A cross-sectional study of rural populations in Bangladesh reported a significant association between general obesity and T2DM. [46] Additionally, a recent geographical study found that several characteristics, such as age, education level, and socioeconomic status(SES) are associated with the increased likelihood of developing HTN and T2DM, with considerable regional differences in Bangladesh. [47] The latest evidence has even shown an inverted U-shaped connection between DM, HTN, and SES. [48] However, active smoking enhances the risk of T2DM, with heavy smokers having the greatest risk, [49] and the risk stays heightened for around ten years after cessation of smoking, declining more rapidly for lighter smokers. [50] In general, gender has long been thought of as a relevant covariate, and [51] there is some indication that exposure to the media has little influence on NCDs.

[52,53,54,55] Becoming noteworthy heretofore by the research studies, there are quite a handful of hypotheses that have been carried forward in an effort to explain the high prevalence of DM and HTN in Bangladesh, including covariates such as gender, older age group, greater educational attainment, better financial status, increased body mass index, unemployment, dwelling territory, and smokers. [56] BDHS 2011 was the first nationwide survey to record fasting blood glucose levels and blood pressure for diagnosing DM and HTN in order to demonstrate the direct relationship with the aforementioned factors and was adopted by the majority of extant research, despite the fact that the data are now very stale. Henceforth, Bangladesh does not have an exhaustive study to assess the determinants of NCDs utilizing among most contemporary data sources (including BDHS 2017-18).

[57]WHO backed the Bangladeshi government in formulating a multisector action plan for the prevention and control of NCDs, a strategy spanning from 2018 to 2025 and including approximately 30 ministries and organizations. As a matter of fact, predicting potential risk factors for NCDs, particularly DM and HTN, is important to initiate rapid public health interventions and deploy more resources. Resultantly, our study focuses on determining the sociodemographic, socioeconomic, and lifestyle-related characteristics that actually affect the incidence of DM and HTN in Bangladesh, in hopes of helping decision-makers prioritize and execute actions to mitigate the financial strain of NCDs to achieve the SDG targets.

## Methods

### Data Source

[58]The research utilized secondary data obtained from the 2017-18 Bangladesh Demographic and Health Survey (BDHS), which is a survey conducted on a nationally representative sample. The 2017-18 Bangladesh Demographic and Health Survey (BDHS) was carried out by the National Institute of Population Research and Training (NIPORT) under the auspices of the Ministry of Health and Family Welfare of Bangladesh, as a component of the broader Demographic and Health Survey (DHS) program. The data was obtained by a two-stage stratified cluster random sampling methodology. The survey’s principal sampling unit (PSU) consists of enumeration areas (EA) that typically encompass an average of approximately 120 homes. In the initial phase, a total of 675 primary sampling units (PSUs) were chosen from a list of 293,579 PSUs generated by the Bangladesh Bureau of Statistics (BBS). The selection of these PSUs was based on a probability proportionate to the size of the enumeration areas (EAs). During the second stage of sampling, a systematic sampling method was employed to choose a total of 30 families from each of the chosen Primary Sampling Units (PSUs).

This study utilizes data from the biomarker questionnaire, which is one of the five types of questionnaires employed in the Bangladesh Demographic and Health Survey (BDHS) conducted in 2017-18. The 2017-18 Bangladesh Demographic and Health Survey (BDHS) is the second iteration of the survey that includes the collection of blood pressure and fasting blood glucose biomarker readings. During the 2017-18 Behavioral Risk Factor Surveillance System (BDHS), data pertaining to biomarkers and pertinent demographic information were gathered from a representative sample of individuals aged 18 and above, residing in one-quarter of the homes chosen for the survey. The collection of biomarkers was undertaken in order to get pertinent national data regarding the prevalence of hypertension (i.e., raised blood pressure) and diabetes (i.e., raised blood glucose). Data regarding the management of hypertension and diabetes was also gathered. The International Classification of Functioning, Disability, and Health (ICF), in collaboration with local experts, provided support in the formulation of the biomarker testing procedure and facilitated the necessary approval from the ICF Institutional Review Board. Prior to their involvement in the study, the participants were required to provide written informed consent. The collection of data was carried out in a confidential manner. Ethical approval is not required for this study as it relies solely on publicly accessible secondary data.

### Sample

In the BDHS 2017-18, data pertaining to biomarkers and pertinent demographic information were gathered from a representative sample of individuals aged 18 and above, residing in one-quarter of the household chosen for participation in the survey. A total of 8,013 females and 6,691 males, all aged 18 years and above, met the criteria for inclusion in the study, which included the assessment of blood pressure and blood glucose levels. Within the surveyed population, it was found that 93% of women and 85% of men underwent blood pressure assessment, whereas 87% of women and 79% of men underwent blood glucose testing.

### Outcome Variable

This study encompasses two primary dependent variables: the presence or absence of diabetes and the presence or absence of hypertension. Individuals were classified as having elevated blood glucose or diabetes if their fasting blood glucose (FBG) exceeded a threshold of 6 mmol/L. Otherwise, they were regarded to be in a normal health status or non-diabetic. Participants were categorized as having hypertension if, during the survey, their mean systolic blood pressure (SBP) exceeded 140 mmHg or their mean diastolic blood pressure (DBP) exceeded 90 mmHg.

### Explanatory Variables

The selection of explanatory variables is informed by a comprehensive evaluation of relevant literature. These variables are further categorized into demographic, biomedical, and behavioral aspects.

The demographic factors considered in this study include age (categorized as <=40 years and >40 years), gender (classified as female and male), division of residence (Barisal, Chittagong, Dhaka, Khulna, Mymensingh, Rajshahi, Rangpur, Sylhet), type of residence (categorized as rural and urban), education level (ranging from no education to primary, secondary, and higher education), and wealth index (categorized as poorest, poorer, middle, richer, and richest).

Biomedical variables encompass body mass index (BMI) categories, including thin (BMI < 18.5), normal (BMI 18.5-24.9), and overweight (BMI > 24.9), as well as the presence of hypertension and diabetes. Diabetes is recognized as a contributing factor to the development of hypertension, while hypertension is similarly acknowledged as a contributing factor to the development of diabetes.

The behavioral characteristics under consideration include smoking status (yes/no), engagement in physical activity or employment (yes/no), and exposure to media (yes/no). An individual is considered to have media exposure if they engage in activities such as reading newspapers or magazines, listening to the radio, or watching television at least once a week.

The categorization process involved adhering to the recommendations provided by the World Health Organization (WHO) and conducting a comprehensive evaluation of relevant literature. As we are using a large survey data, small number of missing values are omitted from the study.

**Figure 1.**
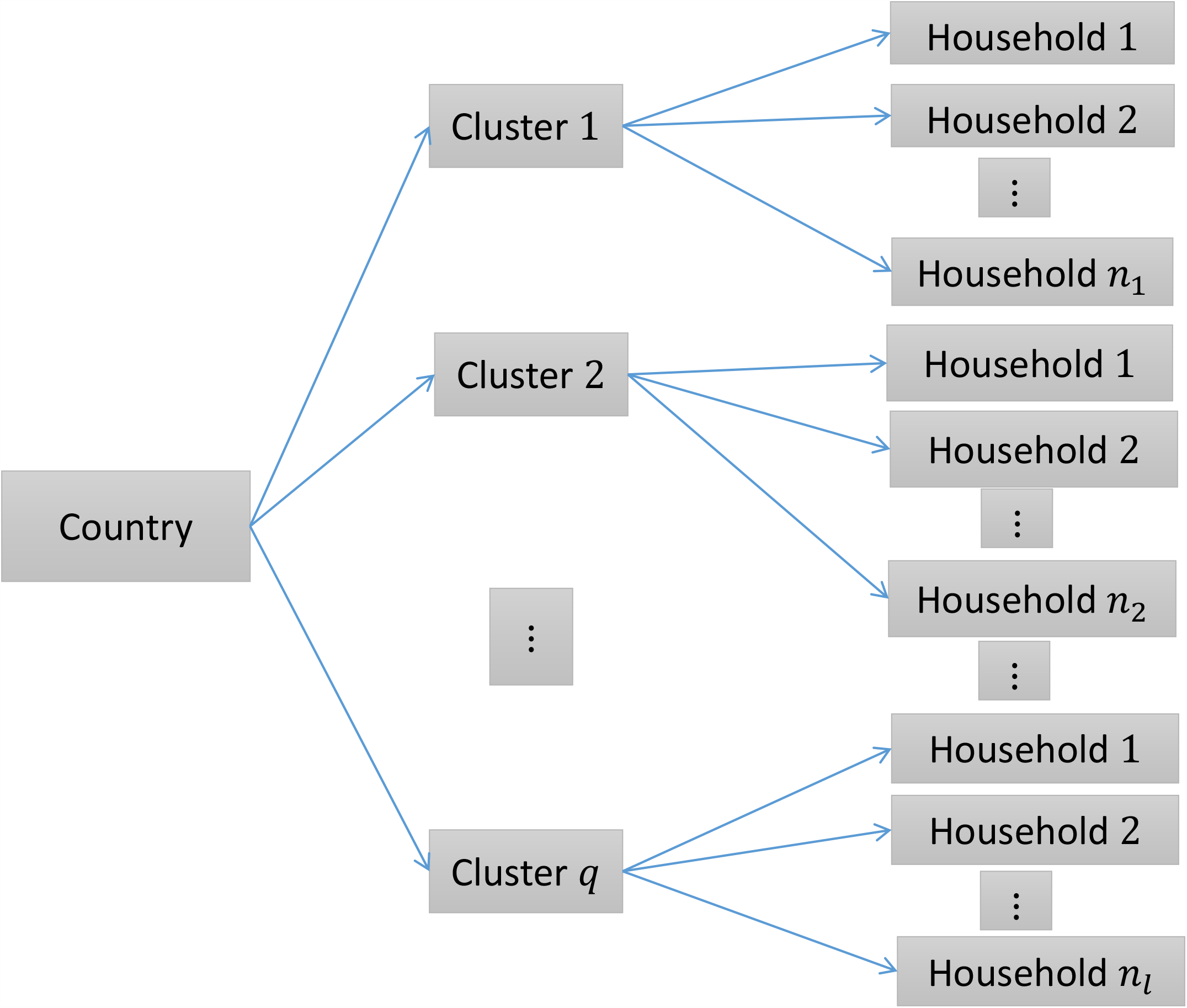
Survey structure

## Why use mixed method

The objective is not only limited to showing the factors of DM and HTN but also to show the clustering effect. A mixed model is required for incorporating the repercussions of the clustering under consideration. For satisfying the intention to encapsulate the clustering effect, in this particular investigation, a mixed-effect logistic regression model was employed as a methodological approach to effectively account for the intricate cluster effects pertaining to the phenomenon of DM and HTN. Through the assessment of intra-cluster correlation (ICC) coefficients, we have ascertained the presence or absence of cluster variation within the dataset employed in the present study.

## Statistical Analysis

In the context of univariate analysis, the frequency distribution of each category within the selected variables is presented in order to illustrate the data patterns across several components. Frequency distributions are utilized to provide a descriptive analysis of the attributes or traits exhibited by the participants or respondents.

In order to investigate the association between several factors and the presence of diabetes and hypertension, a bivariate analysis was undertaken, accompanied by the utilization of the Chi-square test. Contingency tables have been utilized to investigate the distribution of diabetes and hypertension among individuals, as well as their relationship with explanatory variables. The chi-square test is conducted to determine the presence of a connection between the presence of diabetes and hypertension while considering unadjusted possible confounders. This analysis is performed on contingency tables where all expected cell frequencies are more than 5.

To explicitly characterize a diverse range of cluster fluctuations, the authors employed generalized linear mixed models (GLMMs). The adjusted odds ratios (AORs) of the covariates were obtained from the mixed-effect logistic regression model. Let, *y*_*lP*_ be *p*^*th*^ individual/household from *l*^*th*^ cluster where *l* = 1, 2, …, *q* and *p* = 1, 2, …, *n*_*l*_; let, *x*_*lP*_ be a vector of covariates for *p*^*th*^ household from *l*^*th*^ cluster related with fixed effect parameter *β. u* represents a (*q* × 1) vector of random effects corresponding to *q* clusters and *z*_*lP*_ is a special vector of size (*q* × 1) which contains all zeros but a 1 at the *l*^*th*^ position; *l* = 1, 2, …, *q*. Here, μ_*lp*_ = *E*(*Y*_*lp*_|μ_*l*_), where μ_*l*_ be the random effect of cluster *l*. Under GLMM the linear predictor takes the form, 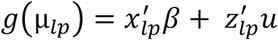. [59] Using the logit link function in GLMM,

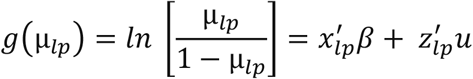

In this study, only random intercept has been considered, 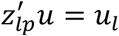, where, 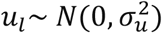. Therefore, the general conditional probability of *p*^*th*^ household from *l*^*th*^ cluster given the value of the explanatory variable of that observation and the random effect of that cluster is,

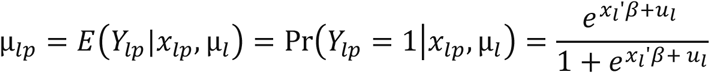

The likelihood function for the individuals corresponding to *l*^*th*^ cluster is,

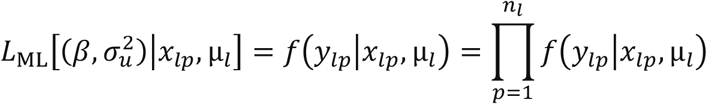

The estimates are obtained by maximizing the following marginal likelihood function,

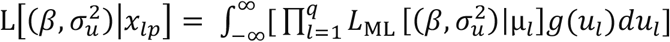

[60]The formula of Intra Cluster Correlation (ICC) coefficient is, 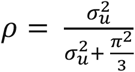

## Results

### Univariate Analysis

We started our analysis by tabulating the frequencies of the variables of interest as shown in Table 1.

**Table 1.** Background Characteristics of the study population.

| **Response Variables** | **Category** | **Count** | **Percent** |
| --- | --- | --- | --- |
| **DIABETES** | Non-Diabetic | 9565 | 78.0% |
|  | Diabetic | 2703 | 22.0% |
| **HYPERTENSION** | Non-Hypertensive | 10170 | 77.5% |
|  | Hypertensive | 2961 | 22.5% |
| **Explanatory Variables** |  |  |  |
| **SEX** | female | 8015 | 54.5% |
|  | male | 6691 | 44.5% |
| **AGE** | <= 40 Years | 8824 | 60.0% |
|  | > 40 Years | 5882 | 40.0% |
| **BMI** | thin | 2203 | 17.1% |
|  | normal | 7519 | 58.2% |
|  | overweight | 3196 | 24.7% |
| **DIVISION** | Dhaka | 2155 | 14.7% |
|  | Barisal | 1541 | 10.5% |
|  | Chattogram | 2072 | 14.1% |
|  | Khulna | 1950 | 13.3% |
|  | Mymensingh | 1644 | 11.2% |
|  | Rajshahi | 1837 | 12.5% |
|  | Rangpur | 1740 | 11.8% |
|  | Sylhet | 1767 | 12.0% |
| **RESIDENCE** | Rural | 9195 | 62.5% |
|  | Urban | 5511 | 37.5% |
| **EDUCATION** | No Education | 3711 | 25.3% |
|  | Primary | 4328 | 29.5% |
|  | Secondary | 4193 | 28.5% |
|  | College or Higher | 2460 | 16.7% |
| **WEALTH INDEX** | Poorest | 2777 | 18.9% |
|  | Poorer | 2664 | 18.1% |
|  | Middle | 2800 | 19.0% |
|  | Richer | 2830 | 19.2% |
|  | Richest | 3635 | 24.7% |
| **MEDIA EXPOSURE** | No | 3030 | 20.6% |
|  | Yes | 11676 | 79.4% |
| **WORKING STATUS** | No | 5909 | 40.2% |
|  | Yes | 8780 | 59.8% |
| **SMOKING STATUS** | No | 11068 | 84.3% |
|  | Yes | 2062 | 15.7% |

Among the participants, 54.5% are male and 44.5% are female. 60% of respondents are less or equal to 40 years old and the other 40% are older than 40 years. More than half of the participants (58.2%) are in the normal category of the BMI measurement scale. It is found that 17.1% are thin and 24.7% are over-weighed. It is observed that 25.3% of participants have no education and 29.5%, 28.5%, and 16.7% of respondents have primary, secondary, and higher education, respectively. Most of the respondents (62.5%) are found to reside in the rural area. The distribution of respondents among the divisions Dhaka, Barisal, Chittagong, Khulna, Rajshahi, Rangpur, and Sylhet are 14.7%, 10.5%,14.1%, 13.3%, 11.2%, 12.5%, 11.8%and 12.0%, respectively. Almost 80% of respondents are exposed to media and 60% are currently working.15.7 % of respondents have a smoking status yes. Around 20% of respondents fall in each category of the wealth index. 22% of respondents are found to be diabetic and 22.5% of respondents have hypertension.

### Bivariate analysis

#### Diabetes

The chi-square test is performed to see the association between having diabetes and Its covariates shown in Table 2. Age, BMI, Hypertension, Division, Residence, Wealth index, Media Exposure, and Working status have significant associations with having diabetes at a 1% level of significance. People are more likely to have diabetes in the age group greater than 40 years (27.1%) than the age group 40 or less than that (18.6%). Diabetes is more prone as BMI Increases as most of the individuals (30.8%) with overweight BMI category have diabetes. People having hypertension are more likely to have diabetes (28.4%) than non-hypertensive patients (20.2%). Dhaka division has the most individuals with diabetes (36.5%) while Rangpur division has the least diabetic patients (14.8%). Urban people are a little bit more prone to have diabetes (23.1%) than rural people (22.2%). Rich people are more likely to have diabetes than poor people according to the percentage of diabetic individuals among the different wealth Categories. Media-exposed people are more likely to have diabetes (23.3%)than people not exposed to media (17.5%) while currently working people are less prone to diabetes (20.4%) than not working people (24.5%). Sex, Education, and smoking status of the individuals were found to have an insignificant association with having diabetes.

**Table 2.**
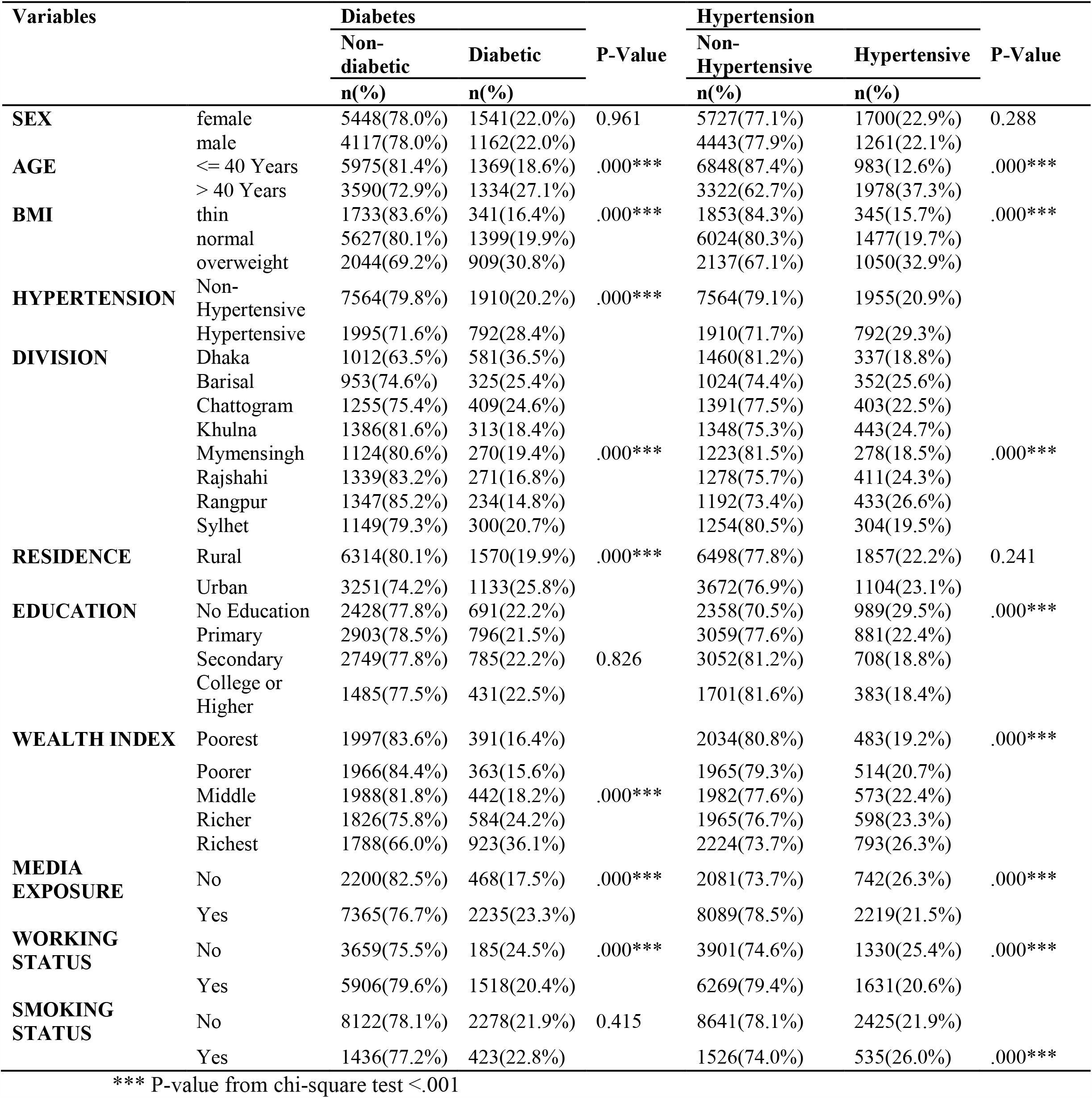
The distribution of Diabetes and Hypertension by sociodemographic variables.

#### Hypertension

The distribution of background characteristics by the levels of hypertension is given in Table 2. Age, BMI, Hypertension, Division, Education, Wealth Index, Media Exposure, Working Status, and Smoking Status have a significant association with having diabetes at a 1% level of significance. Like diabetes, people are more likely to have diabetes in the age group greater than 40 years (37.3%) than the age group 40 or less than that (12.6%). Hypertension is more prone as

BMI Increases as most of the individuals (32.9%) with overweight BMI category have hypertension. People having diabetes are more likely to have hypertension (29.3%) than non-diabetic patients (20.9%). unlike diabetes, the Dhaka division has the least individuals with hypertension (18.6%) while the Rangpur division has the most hypertensive patients (26.6%). Exactly like in the case of diabetes, Urban people are a little bit more prone to have hypertension (23.1%) than rural people (22.2%). Education is negatively associated with hypertension as no educated people have the most hypertensive patients (29.5%) and it reduces as the level of education increases. Rich people are more likely to have hypertension than poor people as the percentage of hypertensive individuals increases with an increase in the wealth index. Media-exposed people are less likely to have hypertension (21.5%)than people not exposed to media (26.3%) also currently working people are less prone to hypertension (20.6%) than not-working people (25.4%). Smoking status has a positive association with hypertension as individuals with smoking are more likely to have hypertension (26.9%) than individuals with no smoking habit (21.9%). Sex and residence were found to have an insignificant association with hypertension.

### Multivariate Analysis

#### Diabetes

The variables that were found to have significant association with complications in bivariate analysis were further examined in a regression analysis for estimating the adjusted effects of these covariates

To determine the potential factors associated with the occurrence of diabetes, adjusted odds ratios obtained from the mixed effect logistic regression model are given in Table 3.

**Table 3.** Adjusted odds ratio (AOR), 95% confidence intervals, and P-value from mixed logistic regression model for diabetes.

| **Variables** | **AOR** | **95%CI** | **P-Value** |
| --- | --- | --- | --- |
| **AGE** |  |  |  |
| ≤40 years | 1 | Reference | - |
| >40 years | 1.748 | 1.58-1.933 | .000*** |
| **BMI** |  |  |  |
| Thin | 0.825 | 0.717-0.95 | .008** |
| Normal | 1 | Reference | - |
| Overweight | 1.455 | 1.302-1.626 | .000*** |
| **Hypertension** |  |  |  |
| Non-Hypertensive | 1 | Reference | - |
| Hypertensive | 1.287 | 1.149-1.442 | .000*** |
| **DIVISION** |  |  |  |
| Dhaka | 1 | Reference | - |
| Barisal | 0.641 | 0.491-0.835 | .000*** |
| Chattogram | 0.544 | 0.426-0.694 | .000*** |
| Khulna | 0.356 | 0.276-0.459 | .000*** |
| Mymensingh | 0.475 | 0.364-0.620 | .000*** |
| Rajshahi | 0.368 | 0.284-0.476 | .000*** |
| Rangpur | 0.315 | 0.241-0.411 | .000*** |
| Sylhet | 0.469 | 0.36-0.611 | .000*** |
| **RESIDENCE** |  |  |  |
| Rural | 1 | Reference | - |
| Urban | 1.018 | 0.876-1.183 | 0.813 |
| **WEALTH INDEX** |  |  |  |
| Poorest | 0.969 | 0.809-1.159 | 0.727 |
| Poorer | 0.878 | 0.740-1.041 | 0.133 |
| Middle | 1 | Reference | - |
| Richer | 1.289 | 1.149-1.536 | .002** |
| Richest | 1.778 | 1.658-2.217 | .000*** |
| **MEDIA EXPOSURE** |  |  |  |
| No | 1 | Reference | - |
| Yes | 1.075 | 0.936-1.234 | 0.306 |
| **WORKING STATUS** |  |  |  |
| No | 1 | Reference | - |
| Yes | 1.015 | 0.919-1.121 | 0.767 |
| Variance Component | 0.38 |  |  |

\*\*\*P-value<.001
\*\*P-value<.01
\*P-value<.05

The odds of having diabetes are 28.7% higher among individuals with hypertension than among individuals not having hypertension as the adjusted odds ratio (AOR) of hypertension is 1.287 with 95% CI (1.149-1.442) for AOR with high statistical significance.

The effect of age with AOR is 1.748 and 95% CI (1.58-1.993) is statistically highly significant for having diabetes i.e. people age greater than 40 years are 74.8% more likely to have diabetes than people aged 40 years or less and these odds could be as low as 1.58 or as much as 1.933.

Individuals in the overweight BMI category have 45.5% higher odds of having diabetes than Individuals with normal weight as the AOR of overweight BMI is 1.455 with a CI (1.302-1.626) for having diabetes which is highly significant. In comparison, individuals in the thin BMI category have 17.5% lower odds of having diabetes than individuals with normal weight as the AOR of thin BMI is 0.825 with CI (0.717-0.95).

Compared to Dhaka division, Barisal, Chattogram, Khulna, Mymensingh, Rajshahi, Rangpur, Sylhet division’s people have 35.9%, 45.6%, 64.4%, 52.5%, 63.2%, 68.5% and 53.1% less odds of having diabetes respectively.

Compared to the people in the middle wealth category, richer people are 28.9% and the richest people are 77.8% more likely to be diabetic patients at a 1% significance level.

#### Hypertension

The adjusted odds ratios obtained by performing mixed logistic regression have been shown in Table 4 to find the risk factors associated with the occurrence of hypertension.

**Table 4.** Adjusted odds ratio (AOR), 95% confidence intervals, and P-value from mixed logistic regression model for hypertension.

| **Variables** | **AOR** | **(95%CI)** | **P-Value** |
| --- | --- | --- | --- |
| **AGE** |  |  |  |
| ≤40 years | 1 | Reference | - |
| >40 years | 4.208 | 3.781-4.685 | .000*** |
| **BMI** |  |  |  |
| Thin | 0.656 | 0.568-0.756 | .000*** |
| Normal | 1 | Reference | - |
| Overweight | 2.154 | 1.929-2.405 | .000*** |
| **DIABETES** |  |  |  |
| Non-Diabetic | 1 | Reference | - |
| Diabetic | 1.244 | 1.112-1.391 | .000*** |
| **DIVISION** |  |  |  |
| Dhaka | 1 | Reference | - |
| Barisal | 1.617 | 1.284-2.036 | .000*** |
| Chattogram | 1.277 | 1.031-1.582 | .025* |
| Khulna | 1.381 | 1.116-1.711 | .003** |
| Mymensingh | 1.099 | 0.871-1.387 | 0.426 |
| Rajshahi | 1.572 | 1.266-1.952 | .000*** |
| Rangpur | 1.927 | 1.548-2.398 | .000*** |
| Sylhet | 1.179 | 0.938-1.481 | 0.157 |
| **EDUCATION** |  |  |  |
| No Education | 1 | Reference | - |
| Primary | 0.931 | 0.821-1.058 | 0.277 |
| Secondary | 0.843 | 0.729-0.974 | 0.021* |
| Higher | 0.749 | 0.625-0.899 | 0.002** |
| **WEALTH INDEX** |  |  |  |
| Poorest | 0.776 | 0.656-.917 | .003** |
| Poorer | 0.842 | 0.719-0.985 | .032* |
| Middle | 1 | Reference | - |
| Richer | 1.081 | 0.929-1.259 | 0.31 |
| Richest | 1.126 | 0.958-1.321 | 0.148 |
| **MEDIA EXPOSURE** |  |  |  |
| No | 1 | Reference | - |
| Yes | 0.811 | 0.713-0.923 | .001** |
| **WORKING STATUS** |  |  |  |
| No | 1 | Reference | - |
| Yes | 0.822 | 0.744-0.908 | .000*** |
| **SMOKING STATUS** |  |  |  |
| No | 1 | Reference | - |
| Yes | 0.902 | 0.792-1.027 | 0.121 |
| Variance Component | 0.126 |  |  |

\*\*\*P-value<.001
\*\*P-value<.01
\*P-value<.05

Having diabetes influences hypertension positively as diabetic patients are 24.4% more likely to have hypertension than non-diabetic people with AOR 1.244 and 95% CI (1.112-1.391).

The adjusted effect of age with AOR=4.208 and 95% CI for AOR (3.781-4.685) is statistically highly significant for having hypertension i.e. people aged greater than 40 years are 3.21 times more likely to have hypertension than people aged 40 years or less.

Increased weight has an impact on hypertension, as the AOR of the overweight BMI category is 2.154 compared to normal weight category individuals. Thus overweight individuals are 2.15 times likely to have hypertension. Whereas thin people have 34.4% lower chances of having hypertension.

Compared to the Dhaka division, Barisal, Chattogram, Khulna, Rajshahi, and Rangpur division’s people have 61.7%, 27.7%, 38.1%, 57.2%, and 92.7% more odds of having hypertension respectively.

Moving from no education to secondary and higher education decreases the odds of having hypertension by 15.7% and 25.1% respectively. Moving from the middle category of the wealth index to the poorer and poorest category decreases the odds of having hypertension by 15.8% and 22.4% respectively.

Media exposure status yes compared to no, reduces the odds of having hypertension by 18.9% with AOR 0.811 and 95% CI (0.713-0.923) at 1 level of significance.

Working people are 17.8% less likely to have hypertension compared to people not working.

The estimates of the variance component of the random effects were found to be 0.38 and 0.126 when modeling diabetes and hypertension respectively. Intra-cluster correlation coefficients from these estimates were found to be 0.103 and 0.369. Both fixed logistic and mixed logistic regression models were applied to analyze the data used in this study and it was found that the Akaike Information Criterion (AIC) value of the mixed model for diabetes was 11760.5 whereas it was 11968.43 for the fixed logistic model. Also, the AIC value from the mixed model for hypertension was 11382.79 and the value was 11409.46 for the fixed model. That is, the mixed model had a smaller AIC value than the fixed model. Therefore, the mixed logistic regression model fits the data better than the fixed logistic regression model. Moreover, the ICC values are 0.104 and 0.037 respectively for the mixed logistic model for DM and HTN, which suggests that there is higher variability across clusters, reinforcing our decision to apply a mixed model to the data.

## Discussion

The main purpose of the study is to find the risk factors for having diabetes and hypertension. To our knowledge, this is the first study to employ a mixed modeling approach to fulfill our purpose. From the mixed effect logistic model, age, BMI, hypertension, division, and wealth index of the rich class are found to have significant effects on diabetes. Diabetes, age, BMI, division, secondary and higher education, wealth index of poorer and poorest, media exposure, and working status significantly affect the likelihood of having hypertension for individuals who are 18 years old or above in Bangladesh.

[61]Hypertension and diabetes share common risk factors and frequently co-occur thus high blood pressure is reported as a significant predictor of diabetes. The study results not only show the significant impact of hypertension on diabetes but also the effect of diabetes on hypertension in Bangladesh.

[64] Previous studies highlight the significance of ongoing monitoring and adequate treatment in light of the rising incidence of type 2 diabetes in senior citizens. [65] Also, it was found increasing age is significantly associated with the increment in blood pressure. Both these findings are also obtained from our analysis.

[65-67] The study found that having an increased level of BMI can enhance the chances of developing T2DM and HTN. This is consistent with previous studies which reported that T2DM and HTN were shown to be substantially associated with increased BMI. At the same time, a BMI of 22.5kg/m2 or more is associated with an increased risk of acquiring diabetes and hypertension in Bangladesh.

[68] Previous studies found that higher economic status can significantly impact the prevalence of diabetes. Our study has also shown a similar result. We found a significant association between the rich class of people and DM, on the other hand, the poorer and the poorest individuals developed hypertension significantly. [69] It is also validated by the previous results as they have shown wealth was strongly related to diabetes but not to hypertension. Apart from these results, in a similar vein, we observed that higher education increases the chances of HTN by 25.1%.

[70] Living in different geographical areas has also a significant impact on both DM and HTN. Interestingly, we found that being able to work and being exposed to the mass media can substantially decrease the chances of HTN, which is also supported by the previous study.

## Strengths and Limitations

One of the strengths of this research is the use of a nationwide large sample with comprehensive information on the occurrence of diabetes and hypertension and related community level and individual-level variables. This data set is collected through a reliable and uniform procedure, which minimizes measurement error and bias. The response rates of this study are high.

The main limitation of this paper is to use of cross-sectional data and hence it may produce selection and information bias. Hence, from this study, it may not be possible to assess the changes in the association between diabetes and hypertension and their risk factors over time. In this study, we only consider respondents of age who are 18 years old or above. Therefore, the results of this study may not be extended to other age groups. Again Diabetes and hypertension are also affected by some other variables such as genetics and family history, tobacco/alcohol consumption, gestational diabetes, polycystic ovary syndrome, unhealthy diet, etc. which are not included in the analysis since these variables are not available in the 2017-18 BDHS data. One may also consider these factors for further analysis.

## Conclusion

[62]The number of individuals with non-communicable diseases is a concerning issue in developing countries such as Bangladesh. Almost 70% of the deaths are caused by complications from non-communicable diseases in Bangladesh. Among many chronic non-communicable diseases, Bangladesh largely suffers from high prevalence of diabetes and hypertension.

[63]Reducing the age threshold for screening females from 40+ years to 35+ years can play a significant role in the detection of diabetes and hypertension consequently providing proper healthcare to the needed one.

The focus should be given to the aforementioned risk factors for having diabetes and hypertension with proper strategy taken by the government of Bangladesh and the health ministry to reduce the prevalence of diabetes and hypertension in Bangladesh.

## Data Availability

All data relevant to the study are available online, and given upon request.
[https://dhsprogram.com/data]

## Acknowledgement

The authors thank the DHS Program for granting access to the BDHS 2017-18 data.

## Data Availability

All data relevant to the study are available online, and given upon request. [https://dhsprogram.com/data]

## Author Contributions

Conception and design: KSAN, TJ; Data curation and formal analysis: KSAN, TJ; Data interpretation: KSAN, TJ; Manuscript draft: KSAN; Reviewed and edited the final manuscript: KSAN.

